# Comparative diagnostic performance of the new chromatographic Affimer^®^-based rapid antigen detection against SARS-CoV-2 and other standard antigen tests for COVID-19 in a clinical setting

**DOI:** 10.1101/2022.01.18.22269401

**Authors:** A.I. Gil-Garcia, A. Lopez-Lopez, J.M. Rubio, J.J. Montoya, Y. Ouahid, A. Madejon, P. Castan

## Abstract

The availability of accurate and rapid diagnostic tools for COVID-19 is essential for tackling the ongoing pandemic. In this context, researchers in the UK have started testing a new Lateral Flow Device (LFD) based on proprietary Biotinylated anti SARS-CoV-2 S1 Affimer® technology that binds to the SARS-CoV2-S1 protein in anterior nasal swab samples, generating an ultra-sensitive method for detection. This international study aimed to compare its performance against other available antigen-detecting rapid diagnostic tests (Ag-RDTs) in a real-world clinical setting. The study was completed under the frame of Project SENSORNAS RTC-20176501 in collaboration with MiRNAX Biosens Ltd. and Hospital Carlos III, it was documented internally and deposited in agreement to the ISO 13485 norm. All the data obtained are currently under submission and review from the Ethics Committee of Universidad Autonoma de Madrid.

## Introduction

Lateral flow devices (LFDs) with sufficient accuracy and ease of use for reliable antigen testing are set to become a cornerstone of SARS-CoV-2 mass community testing. Unluckily, the reduced sensitivity reported for most immunohistochemistry (IHC) methods such as antigen capture enzyme-linked immunosorbent (ELISA) and immunofluorescence assays compared with PCR, has raised questions about reliability and the real performance for identification of infectious cases. Luckily, the development of novel streamlined LFDs which incorporate innovative solutions has increased both, accuracy and fitness for use in the identification of individuals in the community with sufficient viral load to increase their likelihood of infecting others (i.e. Ct ≤30). Obviously, higher Cts, or lower viral loads than this can still be infectious if people are confined in small spaces for long periods or are intimate but reaching the aforementioned threshold about Ct ≤30 would provide a valuable tool to identify most infectious individuals in the community setting.

In this scenario, propelled by the global pandemic, a novel biotinylated anti SARS-CoV-2 S1 Affimer® technology that binds to the SARS-CoV2 S1 protein was characterised in vitro and had its diagnostic accuracy clinically evaluated in late 2021^1^. The system operates generating a complex that migrates along a lateral flow strip by capillary action implementing an innovative ultrasensitive Lateral Flow Device (AffiDX® SARS-CoV-2 Antigen Rapid test - LFD) based on the interaction of immobilized poly-streptavidin with the migrating complex, via the available biotin label on the anti SARS-CoV-2 S1 Affimer®^2,3^. This novel anti SARS-CoV-2 S1 Affimer® technology capitalizes on the scientific confirmation that the SARS-CoV-2 S1 protein binds to the angiotensin converting enzyme 2 (ACE2) receptor in humans mediating fusion of the viral and cellular membranes. Therefore, detection of the SARS-CoV-2 S1 protein in the system is achieved thorough binding to the novel Affimer® technology to the S protein trimer spikes protruding from the viral envelope that bring the viral and cellular membranes close for fusion^4,5^.

The clinical performance of the AffiDX® SARS-CoV-2 Antigen Rapid was initially assessed in the study conducted at different investigational sites in Madrid, with consenting patients of any age, gender, or race/ethnicity who presented at the test site with a former PCR result for COVID19 no older than 4 days^1^. In that study, RTqPCR data were the standard for comparison of the results from 150 nasal swabs taken on the LFD with the AffiDX® SARSCoV-2 Antigen Rapid test.

After analyzing the clinical results obtained in that study with the AffiDX® SARS-CoV-2 Antigen LFD (where a sensitivity of 98% and a specificity of 100% within the cohort tested for a Ct threshold ≤30 was reported), comparison with other commercially available LFDs for detection of SARS-CoV2 antigens was performed within the same framework. This work reports on the data obtained for that comparative study including Abbott Panbio™ COVID-19 Ag Rapid Test Device–(Nasopharyngeal), Innova SARS-CoV-2 Rapid Antigen Test and Biozek Nasal swab -COVID-19-Antigen rapid test cassette systems.

## Objectives

This work aimed to evaluate three commercially available LFDs and the novel Biotinylated anti SARS-CoV-2 S1 Affimer® technology that binds to the SARS-CoV2 S1 protein in anterior nasal swab samples^1^, assessing their correlation with the infectious viral load estimated from PCR cycle threshold (Ct) values. The three LFDs set for comparison were selected through a desk-top review, including manufacturers’ claimed performance and instructions for use, to identify the three commercial tests which, prima facie, may perform with sufficient sensitivity and very high specificity to be fit for use in direct responses to emerging outbreaks.

## Methods

In this retrospective analysis, the diagnostic performance of the new anti SARS-CoV-2 S1 Affimer® technology^1^ and 3 commercial antigen-based LFDs (i.e. Abbott Panbio™ COVID-19 Ag Rapid Test Device (Nasopharyngeal), Innova SARS-CoV-2 Rapid Antigen Test and Biozek Nasal swab -COVID-19-Antigen rapid test cassette) was compared with real-time reverse transcription quantitative polymerase chain reaction assay in terms of sensitivity, specificity and expected predictive values.

The estimation of the clinical performance with clinical samples from volunteers enrolled in test-field epidemiologic studies was designed so as to retrospectively select positive donors who would donate 4 specimens -one for each of the systems compared-. The donors were called back within 48 hours of the PCR confirmation for active SARS-CoV-2 infection, providing one anterior nasal swab and three nasopharyngeal samples for the aforementioned systems according to the manufacturer’s instructions. LFD tests were performed as per the Instructions for Use provided with each product, by minimally trained operators with little laboratory experience who received no previous training on use of the systems and were, therefore, representative of the intended users. LFD results were collected and compared with the PCR results that had been previously obtained, which were set as the gold standard for detection of SARS-CoV-2. The study was completed under the frame of Project SENSORNAS RTC-20176501 in collaboration with MiRNAX Biosens Ltd. and Hospital Carlos III, including a total of 100 samples (50 negatives and 50 positives), each test was documented internally and deposited in agreement to the ISO 13485 norm. All the data obtained under the frame of Project SENSORNAS RTC-20176501 in collaboration with MiRNAX Biosens Ltd. are currently under submission and review from the Ethics Committee of Universidad Autonoma de Madrid. The clinical performance for each LFD was estimated within this retrospective structure test-field study collating the data from the consenting patients of any age, gender, or race/ethnicity who had presented at the test site with a former PCR confirmation for COVID-19 no older than 2 days. Negative specimens were obtained from consenting patients of any age, gender, or race/ethnicity who presented at the test site with a former negative PCR for COVID-19 no older than 4 days. The data were obtained during the time period between March–July 2021 so no inference to the Omicron variant should be done as it wasn’t until November 2021 that the WHO’s Technical Advisory Group on SARS-CoV-2 Virus Evolution declared PANGO lineage B.1.1.529 the variant of concern designated Omicron^6^.

### Study samples

50 positive (with Ct values ≤30) and 50 negative donors all recently confirmed by RTqPCR were asked to provide the anterior nasal swab which was used to assess the AffiDX® antigen LFD and three nasopharyngeal swabs that which were used for the Abbott Panbio Rapid Antigenic CoronaTest -Nasopharyngeal-, Innova SARS-CoV-2 Rapid Antigen Test and Biozek Nasal swab -COVID-19-Antigen rapid test cassette, respectively.

**Table 1.** Summary of consumables used for PCR confirmation from the volunteers enrolled in test-field epidemiologic studies.

|  |  |
| --- | --- |
| Type of swabs | Beckton Dickenson BBL™ CultureSwab™ dry swab |
| Type of extraction | Extraction protocol - Maxwell® RSC Buccal Swab RNA Kit |
| Type of RTqPCR assay | Applied Biosystems TaqPath COVID-19 CE-IVD RT-PCR |

## Results

The clinical performance of the AffiDX® SARS-CoV-2 Antigen Rapid test based on the novel SARS-CoV-2 S1 Affimer® technology and the three commercially available LFD’s (Abbott Panbio™ COVID-19 Ag Rapid Test DeviceInnova SARS-CoV-2 Rapid Antigen Test and Biozek Nasal swab -COVID-19-Antigen rapid test cassette) was evaluated in the study conducted at different investigational sites in Madrid, Spain as it has been detailed in Materials and Methods. Consenting patients of any age, gender, or race/ethnicity who presented at the test site with a former PCR result for COVID-19 no older than 2 days from a positive result and 4 days for a negative were tested. RTqPCR data were the standard for comparison of the results from the nasal swab on the LFD with the AffiDX® SARS-CoV-2 Antigen Rapid test based and the nasopharyngeal swabs on the Abbott Panbio™ COVID-19 Ag Rapid Test Device, Innova SARS-CoV-2 Rapid Antigen Test and Biozek Nasal swab -COVID-19-Antigen rapid test cassette systems.

### Negative cohort

50 nasal samples were identified as negative with the AffiDX® SARS-CoV-2 Antigen LFD matching their former 50 negative RTqPCR results. Therefore, there was 100% correlation for the detection of negative samples as shown in table 2, with no false positives observed.

**Table 2.** Correlation between RTqPCR and the SARS-CoV-2 Antigen LFD (Avacta® AffiDX® test) for the negative cohort.

| Determination | Number of Negative identifications | Specificity |
| --- | --- | --- |
| PCR | 50 |  |
| AffiDX® Antigen - Nasal | 50 | 100% |

50 nasopharyngeal samples were identified as negative with the Abbott Panbio™ COVID-19 Ag Rapid Test Device LFD matching their former 50 negative RTqPCR results. Therefore, there was 100% correlation for the detection of negative samples as shown in table 2, with no false positives observed.

**Table 3.** Correlation between RTqPCR and the Abbott Panbio™ COVID-19 Ag Rapid Test Device LFD for the negative cohort.

| Determination | Number of Negative identifications | Specificity |
| --- | --- | --- |
| PCR | 50 |  |
| Abbott Panbio™ COVID-19 Ag Rapid Test Device LFD | 50 | 100% |

50 nasopharyngeal samples were identified as negative with the Innova SARS-CoV-2 Rapid Antigen Test matching their former 50 negative RTqPCR results. Therefore, there was 100% correlation for the detection of negative samples as shown in table 2, with no false positives observed.

**Table 4.** Correlation between RTqPCR and the Innova SARS-CoV-2 Rapid Antigen Test for the negative cohort.

| Determination | Number of Negative identifications | Specificity |
| --- | --- | --- |
| PCR | 50 |  |
| Innova SARS-CoV-2 Rapid Antigen Test | 50 | 100% |

49 nasopharyngeal samples were identified as negative with the Biozek Nasal swab -COVID-19-Antigen rapid test cassette matching all but one of the former negative RTqPCR results. Therefore, there was 98% correlation for the detection of negative samples as shown in table 2, with one false positive observed.

**Table 5.**
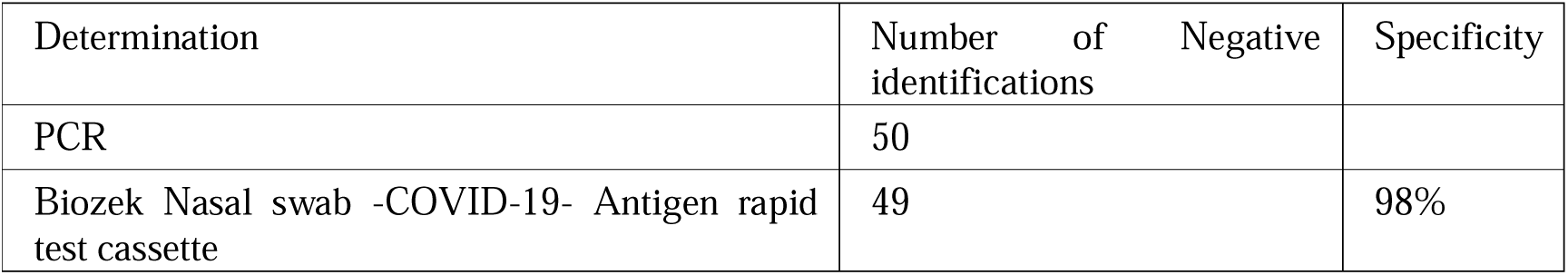
Correlation between RTqPCR and the Biozek Nasal swab -COVID-19- Antigen rapid test cassette for the negative cohort.

### Positive cohort

50 nasal samples were identified as positive with the AffiDX® SARS-CoV-2 Antigen LFD matching their former 50 positive RTqPCR results. Therefore, there was 100% correlation for the detection of positive samples as shown in table 2, with no false negatives observed.

**Table 6.** Correlation between RTqPCR and the SARS-CoV-2 Antigen LFD (Avacta® AffiDX® test) for the positive cohort.

| Determination | Number of Positive identifications | Specificity |
| --- | --- | --- |
| PCR | 50 |  |
| AffiDX® Antigen - Nasal | 50 | 100% |

48 nasopharyngeal samples were identified as positive with the Abbott Panbio™ COVID-19 Ag Rapid Test Device LFD matching all but two of the former 50 positive RTqPCR results. Therefore, there was 96% correlation for the detection of positive samples as shown in table 2, with two false negatives observed.

**Table 7.** Correlation between RTqPCR and the Abbott Panbio™ COVID-19 Ag Rapid Test Device LFD for the positive cohort.

| Determination | Number of Positive identifications | Specificity |
| --- | --- | --- |
| PCR | 50 |  |
| Abbott Panbio™ COVID-19 Ag Rapid Test Device LFD | 48 | 96% |

47 nasopharyngeal samples were identified as positive with the Innova SARS-CoV-2 Rapid Antigen Test matching all but three of the former 50 positive RTqPCR results. Therefore, there was 94% correlation for the detection of positive samples as shown in table 2, with three false negatives observed.

**Table 8.** Correlation between RTqPCR and the Innova SARS-CoV-2 Rapid Antigen Test for the positive cohort.

| Determination | Number of Positive identifications | Specificity |
| --- | --- | --- |
| PCR | 50 |  |
| Innova SARS-CoV-2 Rapid Antigen Test | 47 | 94% |

47 nasal samples were identified as positive with the Biozek Nasal swab -COVID-19-Antigen rapid test cassette matching all but three of the former positive RTqPCR results. Therefore, there was 94% correlation for the detection of negative samples as shown in table 2, with three false negatives observed.

**Table 9.** Correlation between RTqPCR and the Biozek Nasal swab -COVID-19- Antigen rapid test cassette for the positive cohort.

| Determination | Number of Positive identifications | Specificity |
| --- | --- | --- |
| PCR | 50 |  |
| Biozek Nasal swab -COVID-19- Antigen rapid test cassette | 47 | 94% |

**Table 10.**
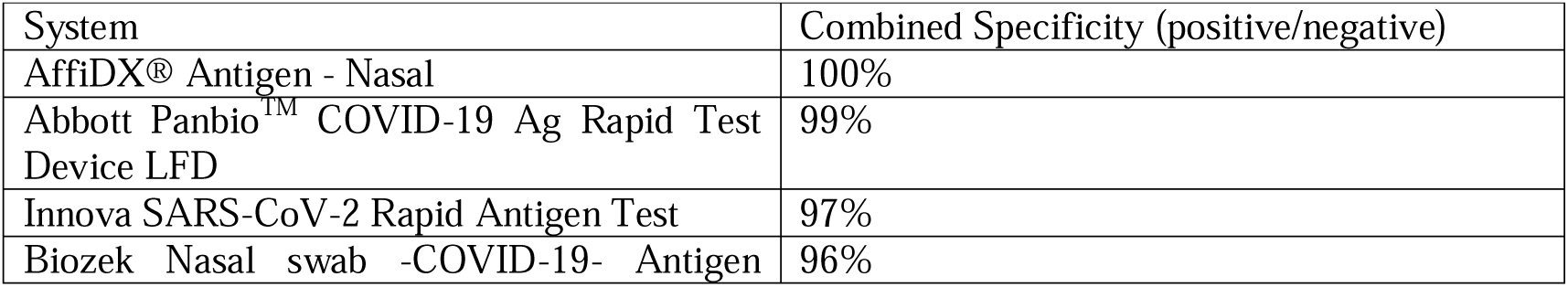
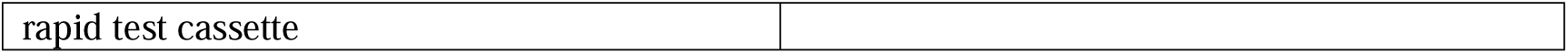
Correlation between RTqPCR and all systems tested.

While all the nasal samples with Ct values of ≤30 were identified as positive with the AffiDX® SARS-CoV-2 Antigen LFD from the cohort of 50 positive RTqPCR results, only 48 were detected with the Abbott Panbio™ COVID-19 Ag Rapid Test Device LFD, and 47 with the Innova SARS-CoV-2 Rapid Antigen Test and the Biozek Nasal swab -COVID-19-Antigen rapid test cassette respectively. Curiously enough, setting a lower Ct threshold for samples with Ct values of ≤28 allowed positive detection for all the 48 specimens within this range for Abbott Panbio™ COVID-19 Ag Rapid Test Device LFD, while Innova SARS-CoV-2 Rapid Antigen Test and Biozek Nasal swab -COVID-19-Antigen rapid test cassette still failed to detect one with a Ct value of 28.00, giving only 47 correct positive results and one false negative each. It was necessary to lower the threshold for samples with Ct values of ≤27 to allow correct determination of the 47 positive specimens within that range for all the systems tested.

**Table 11.**
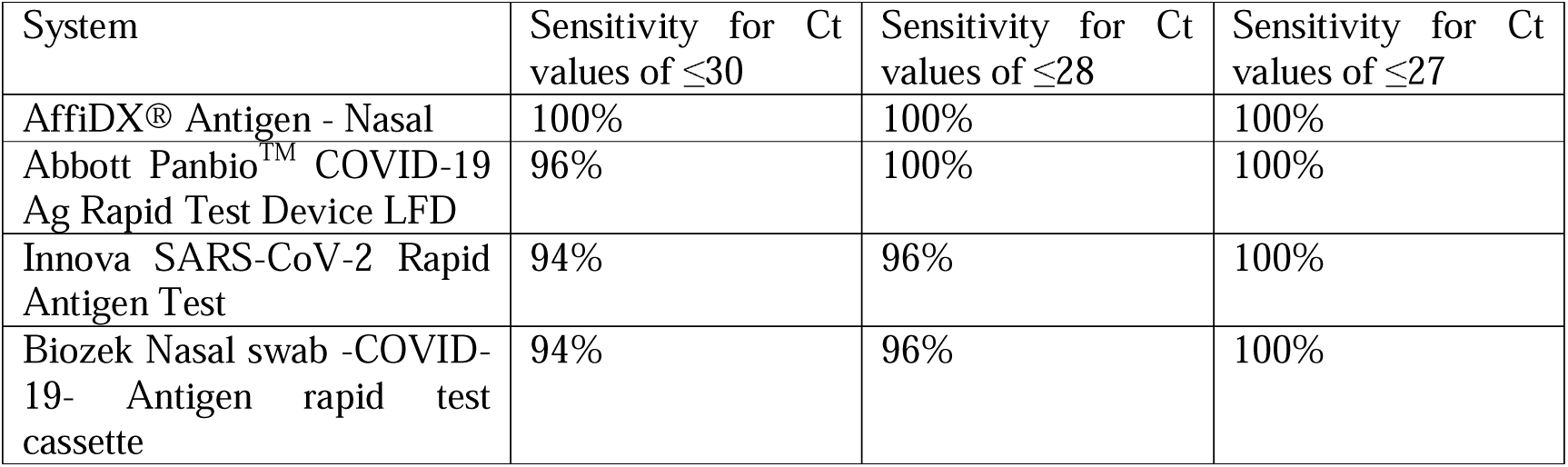
Sensitivity for all systems tested within the three Ct thresholds defined (≤30, ≤28, ≤27).

| System | Sensitivity for Ct values of $\leq 30$ | Sensitivity for Ct values of $\leq 28$ | Sensitivity for Ct values of $\leq 27$ |
| --- | --- | --- | --- |
| AffiDX® Antigen - Nasal | 100% | 100% | 100% |
| Abbott Panbio™ COVID-19 Ag Rapid Test Device LFD | 96% | 100% | 100% |
| Innova SARS-CoV-2 Rapid Antigen Test | 94% | 96% | 100% |
| Biozek Nasal swab -COVID-19- Antigen rapid test cassette | 94% | 96% | 100% |

## Conclusions

The routine use of Ag-RDTs may be convenient in moderate-to-high intensity settings when high volumes of specimens are tested every day. However, the diagnostic performance of the commercially available tests may differ substantially when low viral loads are to be detected. In such scenario, the optimal Ct cut-off value of the new anti SARS-CoV-2 S1 Affimer® technology that maximized sensitivity was 29; improving by (2-3) Ct’s the cut-off value of the 2 Ag-RDT tested, which translates to a 4 to 8 fold increase in sensitivity for viral load detection.’. This result is especially interesting as the 3 commercial antigen-based LFDs (i.e. Abbott Panbio™ COVID-19 Ag Rapid Test Device, Innova SARS-CoV-2 Rapid Antigen Test and Biozek Nasal swab -COVID-19-Antigen rapid test cassette) compared have shown to require higher viral counts to work as effectively.

In summary, the new anti SARS-CoV-2 S1 Affimer® technology has shown to improve the performance at lower viral loads by 2-3 Ct values compared with the 3 other antigen-based LFDs showing fitness for routine use to reduce infections when used in moderate-to-high intensity settings where high volumes of specimens are tested every day.

In this context, the questions raised by some national health authorities about the reliability of these tests, would be answered up to a certain extent by the new AffiDX® SARS-CoV-2 Antigen LFD as it has shown to require a lower viral count (confirmed by RTqPCR) to show positive within the cohort tested.

## Data Availability

The study was completed under the frame of Project SENSORNAS RTC-20176501 in collaboration with MiRNAX Biosens Ltd. and Hospital Carlos III, including a total of 100 samples (50 negatives and 50 positives), each test was documented internally and deposited in agreement to the ISO 13485 norm.

## Acknowledgements

The authors would like to than Dr. Carlos Toro Rueda (Dpt. Of Microbiology of Hospital Universitario la PAZ, Spain), for the unbiased review and amend of all data and text within the paper.

## References

1. Montoya, J.J., Rubio, J.M., Ouahid, Y., Lopez, A., Madejon, A., Gil-Garcia, A.I., Hannam, R.F., Butler, H.R.E. and Castan, P. In vitro characterisation and clinical evaluation of the diagnostic accuracy of a new antigen test for SARS-CoV-2 detection. Journal of Biology and Today’s World 2021, Vol.10, Issue 6, 001–005

2. Dinnes J, Deeks JJ, Adriano A, Berhane S, Davenport C, Dittrich S, et al. Rapid, point-of-care antigen and molecular-based tests for diagnosis of SARS-CoV-2 infection. Cochrane Database Syst Rev. 2020;8(8):CD013705. pmid:32845525

3. Woodman R, Yeh JT, Laurenson S, Ko Ferrigno P (October 2005). “Design and validation of a neutral protein scaffold for the presentation of peptide aptamers”. Journal of Molecular Biology. 352 (5): 1118–33. doi:10.1016/j.jmb.2005.08.001. PMID 16139842.

4. Hoffmann T, Stadler LK, Busby M, Song Q, Buxton AT, Wagner SD, Davis JJ, Ko Ferrigno P (May 2010). “Structure-function studies of an engineered scaffold protein derived from stefin A. I: Development of the SQM variant”. Protein Engineering, Design & Selection. 23 (5): 403– 13. doi:10.1093/protein/gzq012. PMC2851446. PMID 20179045.

5. Stadler LK, Hoffmann T, Tomlinson DC, Song Q, Lee T, Busby M, Nyathi Y, Gendra E, Tiede C, Flanagan K, Cockell SJ, Wipat A, Harwood C, Wagner SD, Knowles MA, Davis JJ, Keegan N, Ferrigno PK (September 2011). “Structure-function studies of an engineered scaffold protein derived from Stefin A. II: Development and applications of the SQT variant”. Protein Engineering, Design & Selection. 24 (9): 751–63. doi:10.1093/protein/gzr019. PMID 21616931.

6. Classification of Omicron (B.1.1.529): SARS-CoV-2 Variant of Concern. World Health Organization. 26 November 2021. Archived from the original on 26 November 2021. Retrieved 26 November 2021.

